# A novel variant in MFN2 linked to a lethal disorder of neonatal onset

**DOI:** 10.1101/2024.09.05.24313021

**Authors:** Mashiat Zaman, Cole Chute, Julien L Marcadier, S Grewal, R Luke Wiseman, Amanda V Tyndall, Brenda McInnes, A Micheil Innes, Francois P Bernier, Timothy E. Shutt

## Abstract

Pathogenic variants in the mitochondrial fusion protein Mitofusin2 typically cause axonal Charcot-Marie-Tooth disease type 2A (CMT2A), a progressively degenerative peripheral neuropathy. Here, we present three siblings with a lethal disorder of neonatal onset who carried a novel homozygous MFN2 variant R334K, which was predicted to be likely pathogenic. Given the severe clinical presentation, which is atypical of MFN2 variants, further functional investigations were warranted to confirm the pathogenicity of the R334K variant. Characterization of patient fibroblasts revealed severe disruptions in all MFN2-related functions assayed, including mitochondrial fragmentation, altered mito-ER contacts, decreased mtDNA copy number and size, increased lipid droplet abundance, and reduced mitochondrial respiration. We also observed reduced Complex I activity, which is noted in cells lacking MFN2, but is not typical of pathogenic MFN2 variants that cause CMT2A. Notably, re-expression of MFN2 R334K in MFN2 knockout cells was unable to rescue mitochondrial fragmentation and the Complex I deficiencies, confirming that the R334K variant is causative of these cellular phenotypes. Furthermore, disease modelling in Drosophila melanogaster demonstrated functional deficiencies of the R334K variant in vivo. Together, these findings confirm that the MFN2 R334K is a novel pathogenic variant causing severe fatal neonatal disease. Finally, we show that pharmacological activation of the integrated stress response rescues the defects caused by MFN2 R334K, offering a potential therapeutic avenue for MFN2 pathology.

## Introduction

Pathogenic variants in MFN2 cause the hereditary motor sensory neuropathy Charcot-Marie-Tooth disease 2a (CMT2a), which is characterized by progressive neuropathy with distal numbness and weakness. CMT2a is generally considered as an early-onset disease, with symptoms typically appearing around age 10. While CMT2a is most often autosomal dominant (CMT2A2A), there are cases of homozygous or compound heterozygous recessive inheritance (CMT2A2B) [25, 46]. MFN2 encodes the integral outer mitochondrial membrane (OMM) protein Mitofusin2 (MFN2), initially characterized for its role in mitochondrial fusion [12]. The protein consists of distinct domains, with an N-terminal GTPase domain, followed by two coiled-coil heptad repeat (HR) domains separated by a transmembrane domain.

The clinical presentation of CMT2a can vary widely, where some MFN2 variants are associated with additional clinical features [29, 53, 54]. While there is evidence of specific MFN2 variants leading to severe and early pathology, neonatal lethality is exceedingly rare [7, 13, 15, 21]. Though the reasons for this heterogeneity are unknown, it is notable that MFN2 has several other functions, including mediating mito-ER contacts (MERCs) [10], facilitating mito-lipid droplet contacts [9], participating in mitochondrial transport [40] and being a key regulator of mitophagy [38]. These MFN2 functions also influence downstream processes such as mtDNA regulation [55], cellular Ca^2+^ signalling [1] and mitochondrial bioenergetics [64]. The impact of different MFN2 variants on the various functions of MFN2 is largely unknown [64]. Surprisingly, for the few variants studied, fusion is not always impacted [2, 16, 30, 33, 63], suggesting that other MFN2 functions are likely contributing to MFN2 pathogenesis.

There is currently no specific therapy for MFN2 pathologies. Although several promising preclinical approaches are in development for CMT2A [49, 50, 62], they focus primarily on promoting mitochondrial fusion, rather than other MFN2 functions. Building on the fact that activation of the Integrated Stress Response (ISR) has recently garnered attention for ameliorating mitochondrial dysfunction [1, 31, 40, 54], we recently showed that ISR activation can restore mitochondrial morphology in patient fibroblasts harbouring the pathogenic D414V MFN2 variant [5], as well as MERCs, mitochondrial motility, and mitochondrial respiration in MFN2 KO cells [8]. ISR activation is mediated by phosphorylation of eIF2α via four kinases that sense different types of cellular stress (i.e., HRI, GCN2, PERK and PRK), ultimately turning on the ATF4 transcription factor that engages multiple cellular stress responses. Thus, ISR activation is a promising therapeutic approach to target mitochondrial dysfunction beyond just impaired mitochondrial fusion.

In this work, we describe three siblings who presented with severe neonatal encephalopathy with areflexia, limited spontaneous movements, respiratory failure, severe hypotonia, arthrogryposis and neonatal demise. All three patients had novel homozygous variants in MFN2, c.1001G>A, p.(Arg334Lys). We report detailed patient phenotypes and provide a comprehensive characterization of multiple mitochondrial functions associated with MFN2 in patient fibroblasts. Using cell and animal modelling, we provide evidence that the R334K MFN2 variant significantly impacts mitochondrial function, not only explaining the cellular phenotypes, but also being responsible for the neonatal lethal axonal disease reported in these three siblings, thus expanding the clinical spectrum of CMT2A. Finally, we show that ISR activation rescues mitochondrial fragmentation and reduced respiration in R334K patient fibroblasts, offering a promising therapeutic avenue for MFN2-mediated pathology.

## Methods

### Genomic analysis

The proband was enrolled into a research study titled Rapid Access for Pediatric Diagnoses Kidomics (RAPiD) at the Alberta Children’s Hospital. Formal written consent was provided by the family (mother, father, and proband) for both the RAPiD research exome (study approved by the University of Calgary Conjoint Health Research Ethics Board) sequencing and the clinical whole exome sequencing. DNA was extracted from whole blood samples, and trio exome library preparation was conducted using the xGen Exome Research Panel v1.0 capture kit (Integrated DNA Technologies, Inc., Coralville, IA, USA). Paired-end sequencing was performed on a NextSeq 550 (Illumina, San Diego, CA, USA) in which data was automatically uploaded into BaseSpace Sequencing Hub where alignment, variant calling, and variant interpretation was completed.

### Sequence alignment

Amino acid sequence alignments for MFN2 and FOXRED1 were performed on T-COFFEE multiple sequence alignment tool [17]. Protein sequences were collected from the UniProt database [58]. The UniProt Sequence ID’s were as follows: Human MFN2: O95140-1; Human MFN1: Q8IWA4; Mouse MFN2: Q80U63; Fish MFN2: A8WIN6; Fly MFN2: Q7YU24; Worm MFN2: Q23424; FXRD1: Q96CU9; FXRD1 [2]: Q96CU9-2 ; FXRD1 [3]: Q96CU9-3; Mouse FXRD1: Q3TQB2; Fish FXRD1: A0A0R4IHD6; Fly FXRD1: Q9VJ10; Worm FXRD1: A0A0V1BI76.

### Cell culture

Skin fibroblasts were cultured in Minimum Essential Media (Gibco™, 11095098), supplemented with 10% Fetal Bovine Serum (FBS) and 10mM Sodium Pyruvate. The cells were maintained at 37°C and 5% CO_2_. U2OS cells were maintained in DMEM high-glucose media (Gibco™; 11965118), also at 37°C and 5% CO_2_. The following ISR inhibitors/activators were used for the morphology and OCR studies: ISRIB (ISR inhibitor) (Cat# SML0843, Sigma), halofuginone (Cat#50-576-3001, Sigma), #3610 or Parogrelil (139145-27-0, Axon Medchem). For drug treatments the compounds were added at the following doses: ISRIB (200nM), compound #3610 (10uM), Parogrelil (10uM) and Halofuginone (100nM),

### Respiration studies

Cellular oxygen consumption rates were measured using the Seahorse XFe24 Extracellular Flux Analyzer (Agilent Technologies) as previously described [52]. In summary, cells were seeded in the manufacturer provided microplate at 70,000 cells/well and incubated for 24 hours at 37°C and 5% CO_2_. Subsequently, the growth media was replaced, and cells equilibrated in assay media supplemented with d-Glucose (25 mM), sodium pyruvate (2 mM) and l-Glutamine (4 mM). Oxygen consumption rates were calculated following administration of the following compounds: Oligomycin (1 μg/mL) (Enzo Life Sciences, BML-CM111), Carbonyl Cyanide 4-(trifluoromethoxy) Phenylhydrazone (FCCP, 1 μM) (Enzo life Sciences, BML-CM120) and Antimycin A (1 μM) (Sigma Aldrich, A8674). The data from the Seahorse analyzer was normalized to protein content data for each well measured by BCA assay (Thermo Fisher Scientific, 23225). The Lucid Resipher assay was used for the ISR studies, as previously described [8]. Briefly, 15,000 cells were seeded on a round bottom 96-well cell culture plate (MaxBioChem, MBC.TC.F. 96)and allowed to settle for 18 hours. The ISR inhibitors/activators were added at the following doses: ISRIB (200nM), compound #3610 (10uM), Parogrelil (10uM) and Halofuginone (100nM):, and OCR was measured at 6 hours after treatment. Samples were normalized for total protein content, lysed from the 96-well plate using RIPA buffer (Thermo Scientific™, 89900) using a BCA assay, standardized with BSA (Fisher Scientific, PI23209).

### Immunofluorescence and Microscopy

20,000 control/patient fibroblasts or U2OS cell lines were seeded on 12mm (Fisherbrand, 12-545-81) coverslips inserted into wells of a 24-well plate (Sarstedt, 83.3922) and incubated for 24 hours at 37°C and 5% CO_2_. Cells were fixed with 4% paraformaldehyde (ElectronMicroscopySciences, 15710) at 37°C for 20 minutes, permeabilized using 0.1% Triton® X-100 (MP Biomedicals, 9002-93-1) and blocked with 10% FBS. The mitochondrial network was stained with anti-TOMM20 (Sigma-Aldrich, HPA011562/Santa Cruz Biotechnology, F-10 sc-17764) primary antibodies and visualized using goat anti-rabbit secondary conjugated with Alexa Fluor 488 (ThermoFisher, A-11034) or goat anti-mouse conjugated Alexa Fluor (ThermoFisher, A-31571). Mitochondrial nucleoids were visualized using anti-dsDNA antibody (Developmental Studies Hybridoma Bank, AB_10805293) and goat anti-mouse secondary conjugated with Alexa fluor 568 (ThermoFisher, A-11004), following permeabilization with 0.2% Triton® X-100). Lipid droplets were studied using BODIPY™ 493/503 (Invitrogen, D3922). Live cells were seeded onto 35mm live cell imaging dishes (Cellvis, D35-20-1.5-N) and allowed to settle for 24 hours. The BODIPY dye was subsequently added at 10uM concentration and incubated for 30 minutes, after which the cells were washed three times with culture media and then imaged. Confocal imaging was performed using Olympus spinning disc confocal. Confocal imaging was performed using Olympus spinning disc confocal system (Olympus SD OSR) (UAPON 100XOTIRF/1.49 oil objective). STED imaging was performed using Leica TauSTED microscope. For the ISR studies, control and patient fibroblasts were seeded at 70,000 cells onto 35mm live cell imaging dishes (D35-20-1.5-N, Cellvis) and allowed to settle for 24 hours. Mitochondrial networks were stained with 100nM MitoTracker Green FM (M7514, Life Technologies) for 30 minutes and subsequently washed with cell culture media three times. . Z-stacks were captured using confocal imaging was performed using Olympus spinning disc confocal system (Olympus SD OSR) (UPlanApo. 60X 1.50NA), including a cellVivo live cell incubator system used to maintain cells at 37°C and 5% CO_2_, Mitochondrial morphology was qualitatively scored based on appearance categorized into fused/intermediate/fragmented for 150 cells, spread across 3 biological replicates. Quantitative mitochondrial network analyses were performed using MiNA on FIJI [59], also scored for 150 cells, spread across 3 biological replicates. Lipid droplets number and size were quantified using the ‘analyze particles’ tool on FIJI.

### Proximity Ligation Assay

The proximity ligation assay was performed as mentioned in [54]. All immunostaining steps, including fixation, blocking and permeabilizing were the same as mentioned for mitochondrial network studies. The Duolink® In Situ Proximity Ligation Assay (Millipore Sigma) was used for this. The aforementioned anti-TOMM20 antibody was used to label the mitochondria, while the ER was labelled using anti-Calnexin (mouse monoclonal, EMD Millipore: MAB3126 (RRID:AB_2069152). Images were analyzed using confocal microscopy (as described above) and PLA puncta were analyzed using the ‘analyze particles’ tool on FIJI.

### Western Blot Analysis

Cells grown to the same confluency were collected and RIPA buffer (Thermo Scientific™, 89900) complimented with protease inhibitors (VWR, 97063-010) was used to lyse and collect the protein extracts. For fly protein, 20 flies were homogenized and sonicated in RIPA buffer (Thermo Scientific™, 89900). Following quantification of protein concentrations using a BCA assay, standardized to BSA (Fisher Scientific, PI23209), 50 μg of total protein was loaded on an SDS-PAGE gel. Subsequently, PVDF membranes (Bio-Rad, 1620177) were used for overnight transfer of the blots. The blots were probed with the following antibodies: anti-MFN2 (Abnova, H00009927-M03; 1:1000), anti-MFN1 (Cell Signaling Technology, D6E2S; 1:1000), anti-OPA1 (BD Transduction Laboratories, 612607; 1:1000), anti-Actin (Sigma, A5316; 1:1000) anti-Actin for fly membrane (DSHB, JLA20; 1:1000) and cocktail anti-OXPHOS Complex antibody (AbCam, ab110411; 1:500). Complimentary horseradish peroxidase conjugated antibodies were used: goat anti rabbit IgG, HRP linked Antibody (Cell Signaling Technology, 7074S) or goat anti-mouse IgG-HRP (Santa Cruz Biotechnology, sc-2055). Blots were treated with the SuperSignal™ West Femto Maximum Sensitivity Substrate (Thermo Scientific™, 34095), and luminescence visualized with an Amersham Imager AI600.

### mtDNA Copy Number and mtDNA Deletions Analysis

Total DNA was extracted and purified from control and patient fibroblasts seeded at 30,000 cells using the PureLink Genomic DNA Mini Kit (Thermo Fisher Scientific, K182001) using the given protocol. Relative mtDNA copy number was analysed using QuantStudio 6 Flex Real-Time PCR system (Thermo Fisher Scientific). The mtDNA and nuclear encoded housekeeping gene 18S were amplified using the primers and reaction conditions described in [18]. Long range PCR reactions were performed to examine mtDNA deletions. The following primers were used for amplification of the mtDNA: 1482–1516 F: ACCGCCCGTCACCCTCCTCAAGTATACTTCAAAGG; 1180–1146 R: ACCGCCAGGTCCTTTGAGTTTTAAGCTGTGGCTCG. The Takara LA Taq Polymerase (Takara Bio, RR002M) was used with 250ng total cellular DNA. PCR cycling conditions were 94°C for 1 min; 98°C for 10 s and 68°C for 11 min (step 2 and 3- 35 cycles); and a final extension cycle at 72°C for 10 min, using an Eppendorf® 5331 MasterCycler Gradient Thermal Cycler. PCR products were visualized by electrophoresis on a 0·6% agarose gel, run for approximately 18 h at 20 V.

### Complex-I Activity Assay

The mitochondrial complex-I activity assay was performed using the Complex I Enzyme Activity Dipstick Assay Kit (AbCam, ab109720) according to manufacturer instructions. Briefly, 50,000 cells were seeded into a six-well plate (Sarstedt, 83.3920) and collected the following day. 10x Extraction buffer from the kit was added to the cell pellet and subsequently spun at 18,000 x g. Supernatants were collected and following homogenization, control and patient samples were plated in the plate provided. Subsequently, the dipstick from the plate was added and incubated for 45 minutes. Following washing procedures, the dipsticks were imaged with Amersham Imager AI600.

### Re-expression of MFN2 R334K in U2OS cells

U2OS MFN2 knockout cells were a generous gift from Dr. Edward Fon (McGill University, Canada) [38]. The MFN2 R334K variant was inserted into MFN2 KO cells as previously described [63]. Briefly, a retroviral plasmid vector (Vector Builder, VB230626-1399nzw) with the MFN2 R334K open reading frame or WT MFN2, followed by an mNeonGreen (after IRES) was virally inserted into MFN2 KO cells, and cells were sorted for mNeonGreen expression through fluorescence activated cell sorting. Endogenous expression levels were quantified by western blot analysis, as described in the western blot section of methods.

### Drosophila stocks, transgenic lines and experiments

All Drosophila were maintained at 25°C and kept in standard food. Flies were allowed to lay eggs on standard grape plates with yeast plated for two days before collection. Drosophila stocks for ey3.5-Gal4 (BDSC#8221) was provided by Dr. William Brook (University of Calgary, Canada), elav-Gal4, tub-Gal4 (BDSC#5138), act5c-Gal4 (BDSC#4414), daGal4, UAS-mCherryRNAi and UAS-marf RNAi (31157) [44] was provided by Dr. Savraj Grewal (University of Calgary, Canada) and tincΔ4Gal4 was provided by Dr. Rolf Bodmer (Sanford Burnham Prebys Medical Discovery Institute, USA). A second UAS-marfRNAi (67158) was purchased from Bloomington Stock Centre (Indiana University, USA). Flies re-expressing the human MFN2 WT and MFN2 R334K were generated through microinjection (GENOMEPROLAB, Canada). The pGW.UAS hMFN2 plasmid used for microinjection was provided by Dr. Paul Marcogliese (University of Manitoba, Canada), which was subsequently mutated by in-fusion cloning (Takara, 638945). Flies were microinjected on chromosome II using the attp40 landing site, through the phiC31 integration approach. For the survival assay, flies were allowed to lay eggs on grape plates with yeast paste for 48 hours and subsequently 50 eggs were collected and placed into standard food. The number of eggs that became pupa were counted and normalized to total number of eggs collected, for percentage survival. For negative geotaxis studies, 30 newly emerged adult flies of the respective crosses were placed into a 1000cc cylinder. On a given day, the flies were gently displaced to the bottom every 3 hours and the percentage of flies that made it past the 10 cm mark after 10 seconds was counted.

### Statistical Analyses

All statistical analyses were performed using GraphPad Prism 9.

## Results

### Case description

#### Proband (P1)

The male proband was born by C-section for preterm labor, breech presentation and fetal heart rate abnormalities at 35 weeks to a 21-25-year-old G3, P0 mother. The pregnancy was remarkable only for a mild COVID infection in the first trimester, with three antenatal ultrasounds reported as completely normal. Decreased fetal movements were noted in the last 4 to 8 weeks of the pregnancy. His birthweight was 2.08 kg (10th percentile), length 46.6 cm (50th percentile) and his head circumference was 32.1 cm (50th percentile). APGARs were 1,3,3,6 at 1, 5,10 and 15 minutes respectively. He remained flat with no respiratory effort and was intubated at 6 minutes of life. He remained ventilator dependent and eventually died in the pediatric intensive care unit at 2-6 weeks of age. On examination, he was noted to have hypotonia both peripherally and axially with decreased spontaneous movements. He had bilateral elbow and knee flexion contracures, with limited hip abduction, and talipes equinovalgus. He was noted to have a bulbous nasal tip, but a complete dysmorphology exam was not feasible.

Investigations included normal newborn metabolic screening, normal rapid aneuploidy detection, SNP chromosomal microarray, Prader Willi, DM-1 and SMA. While chromosomal microarray analysis did not reveal any copy number variation to explain the proband’s phenotype, the SNP component of the analysis revealed three contiguous regions (greater than 10Mb) of absence of heterozygosity (AOH) within chromosomes 1, 3, and 4. Metabolic investigations revealed normal urine and plasma amino acids and a normal plasma acylcarnitine profile. Urine organic acids were non diagnostic and plasma 3-methylglutaconic acid and 3-methylglutaric acid were also normal. EEG was indicative of moderate diffuse/multifocal cerebral dysfunction with multifocal epileptiform discharges. Brain MRI shortly after birth showed multiple foci of diffusion restriction due to cytotoxic edema involving the deep and periventricular white matter in frontoparietal lobes with corresponding intrinsic T1 hyperintensity. There were no overt signs on imaging or MRS to suggest a metabolic cause. A follow-up brain MRI just prior to death reported interval resolution of the multiple foci of restricted diffusion in the bilateral centrum semiovale ovale/corona radiata, with a few small scattered residual centrum semiovale T1 hyperintense foci without restricted diffusion. As compared to the previous MRI, there was interval development of atrophy of the superior vermis, seen as increased interfoliate distance. Mild atrophy was also seen involving the lateral cerebellar hemispheres. The brain stem structures, especially the midbrain and pons, had reduced volume for patient’s age, with prominence of the prepontine cisterns, resulting in prominence of the posterior fossa CSF spaces without mass effect. The remainder of the midline structures appeared normal with no Chiari I deformity and a normal pituitary gland. The corpus callosum was well developed and the remaining brain parenchyma was normal with an age-appropriate myelination pattern. No new areas of restricted diffusion or abnormal signal involving the cerebrum or cerebellum were observed and there was no abnormal susceptibility-related signal loss to suggest blood degradation products. The brain stem structures, especially the midbrain and pons had reduced volume for the patient’s age. Additional imaging of the patient’s spine showed marked smooth hypoplasia and reduced caliber of the thoracic spinal cord. In addition, the brainstem and thoracic spinal cord reported to be small.

A muscle biopsy was performed shortly before death revealing no definite abnormalities but coarse NADH, SDH, and COX histochemistries were observed in addition to mild mitochondrial aggregation in the subarcolemmal spaces with some smaller mitochondria but normal cristae. There were grouped atrophic fibres and occasional hypertrophic fibres.

He was born to healthy Middle Eastern parents who were known to be distantly consanguineous. There was no family history of similarly affected children.

#### Proband’s sibling (P2)

The proband’s male sibling presented with the same core phenotype of encephalopathy, hypotonia, hyporeflexia, arthrogryposis. The sibling had an unremarkable prenatal history with normal anatomy scan. A second scan at revealed breech presentation but otherwise normal with normal fetal movements. A further scan was appropriate for gestational age and showed breech presentation. He was born via C-section due to preterm labor with APGARS 1, 2, and 3 at 1, 5 and 10 minutes respectively, with no spontaneous respirations. He was intubated and admitted to NICU. He required continuous mandatory ventilation. Life sustaining measures were withdrawn between day 11-15 and the patient died the same day. An autopsy was declined.

On examination at birth, the baby was noted to be hypotonic with contractures and decreased range of motion of elbows, hips and knees with bilateral talipes equinovarus. His birth weight was 2.365kg (<50^th^ percentile), length 48 cm (<90^th^ percentile) and head circumference 31.4 cm (50^th^ percentile). He had depressed/absent reflexes. Investigations included a normal newborn metabolic screen, normal lactates, ammonia, and liver enzymes with a mildly elevated CK. An EEG was consistent with encephalopathy. Brain MRI showed cerebellar hypoplasia with query germiolytic cysts. An MRI of the spine was normal.

#### Proband’s sibling (P3)

A third affected child presented with very similar features to other affected siblings. Prenatal ultrasounds at 23-27 weeks revealed suspected club feet, and the fetus was in breech presentation at 32-36 weeks, which raised concerns regarding a likely recurrence of what was now clearly suspected to be a recessive disorder. Delivery was by emergent C-section with APGARS of 1,2,4. The neonatal presentation was very similar to the siblings, with severe encephalopathy, limited respiratory effort, no large joint spontaneous movements or spontaneous eye opening, and arthrogryposis requiring admission to NICU for ventilation. No seizures were identified, and the EEG was encephalopathic with prolonged interburst intervals. MRI features were also consistent with those of the previous affected children with hypoplasia of the parietal lobes, cerebellum, and upper cord. The goal of care was changed at 3-7 days of life, and this girl passed shortly after extubation.

### Genetic workup

Upon initial research RAPID exome sequencing, trio analysis of P1 and parents, who were distantly consanguineous, detected the homozygous c.1001G>A, p.(Arg334Lys) variant of uncertain significance in the MFN2 gene. Pathogenic variants in MFN2 are associated with autosomal dominant and autosomal recessive Charcot-Marie-Tooth disease, axonal, type 2A. While patients with variants in MFN2 have a heterogeneous clinical presentation with an age of onset ranging from 1-45 years, previously reported individuals with homozygous or compound heterozygous variants presented before the age of 10 years [41, 57]. These affected individuals presented with muscle weakness, distal sensory loss, and normal or mildly decreased nerve conduction velocity, while the heterozygous parents were reported to be asymptomatic or had signs of a mild peripheral neuropathy [41]. This variant has not been reported in gnomAD, ClinVar, or the literature. This missense variant resides near the middle of exon 10 of 19 and appears to abolish a potential cryptic donor splice site within this exon. The consequence of the loss of this cryptic donor splice site is unknown. A subsequent clinical exome of P1 confirmed the homozygous MFN2 c.1001G>A, p.(Arg334Lys) variant, and also found a novel homozygous missense variant of unknown significance, c.1454T>A, p.(Ile485Asn), in FOXRED1, a gene associated with complex I deficiency [4, 20, 22, 31].

Sanger sequencing was used to assess the genetic status of P2. Analysis found homozygosity for MFN2 Arg334Lys, but only heterozygosity for FOXRED1 Ile485Asn. The family also had an unaffected sister, for whom no genetic information was obtained.

Based on the genetic work-up from P1, we initially considered both MFN2 R334K and FOXRED1 I485N variants. Given that the candidate variants in MFN2 and FOXRED1 have not been described or characterized previously, we examined the conservation of the corresponding amino acid changes in the protein. We found that the MFN2 R334 residue is conserved in multiple species, including fruit flies and worms, as well as in the paralogous protein MFN1, arguing that it plays an important role in protein function (Figure 1a). Meanwhile, though the FOXRED1 I485 residue is conserved in vertebrates, it is not conserved in earlier diverging metazoans such as flies or worms (Figure 1b). Upon subsequent sequencing of P2, who was homozygous for MFN2 R334K, but only heterozygous for and FOXRED1 I485N, it became evident that MFN2 R334K was most likely the causative variant for the remarkably consistent phenotype in the two brothers. Topologically, the MFN2 R334K variant is near the GTPase domain (Figure 1c), which is crucial for mitochondrial fusion [32].

**Figure 1:**
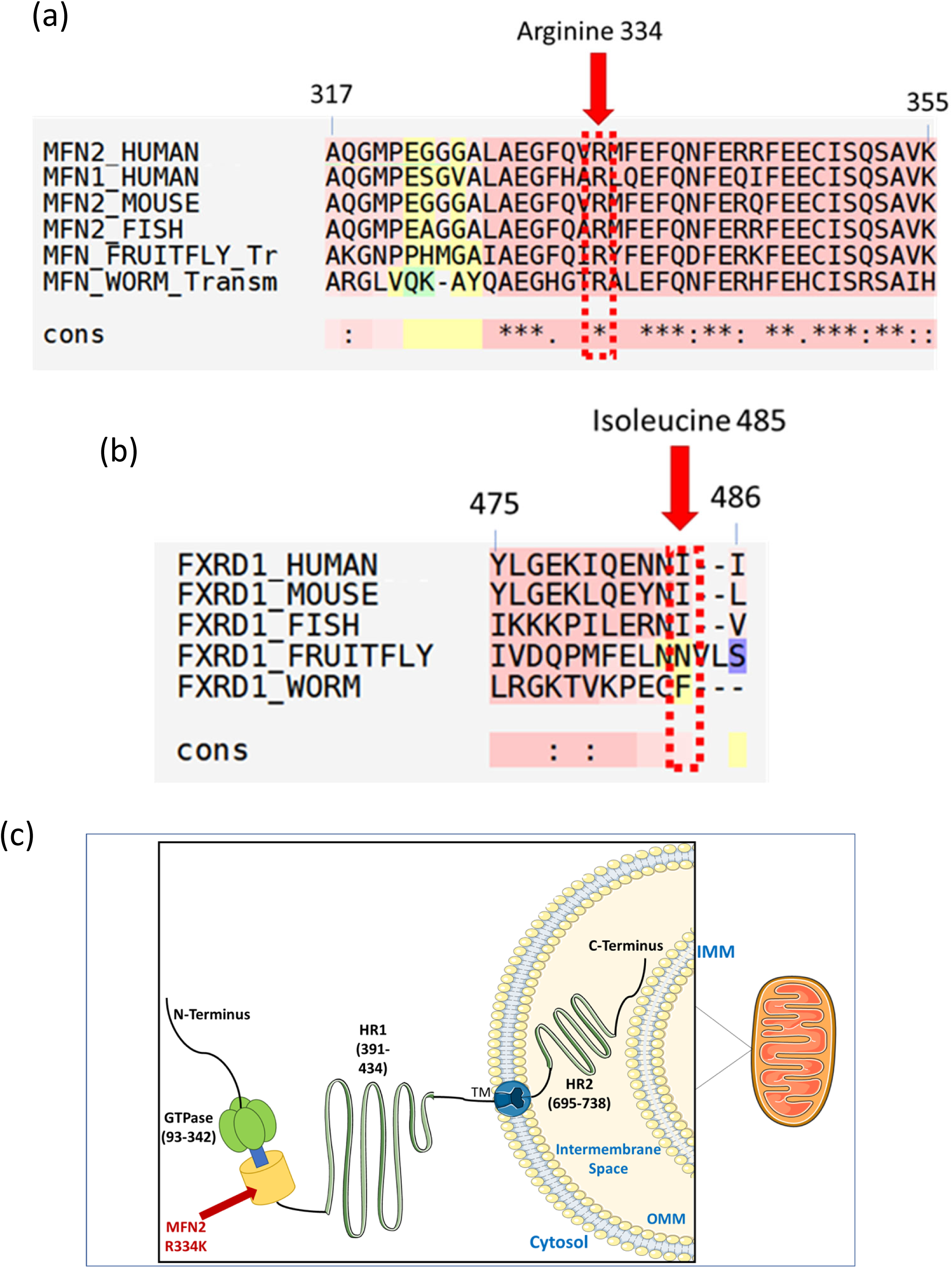
The MFN2 and FOXRED1 single nucleotide variants are conserved across species. (a) Multiple sequence alignment data shows conservation of MFN2 at Arginine-334 and (b) FOXRED1 conservation of Isoleucine 485. Accession numbers used for comparison are: MFN2: O95140-1; O95140-2; MFN1: Q8IWA4; Mouse MFN2: Q80U63; Fish MFN2: A8WIN6; Fly MFN2: Q7YU24; Worm MFN2: Q23424; FXRD1: Q96CU9; FXRD1 [2]: Q96CU9-2; FXRD1 [3]: Q96CU9-3; Mouse FXRD1: Q3TQB2; Fish FXRD1: A0A0R4IHD6; Fly FXRD1: Q9VJ10; Worm FXRD1: A0A0V1BI76. (c) Topological representation of Mitofusin 2, highlighting the position of MFN2 R334K on the GTPase domain.

### Characterization of patient fibroblasts harboring MFN2 R334K

As the candidate pathogenic variant in MFN2 is novel and has not been characterized previously, fibroblasts from a skin biopsy of P1, who harboured homozygous MFN2 R334K and FOXRED1 I485N variants, were obtained for functional analysis. We examined a variety of cellular and mitochondrial phenotypes to see which, if any, might be consistent with MFN2 and/or FOXRED1 dysfunction. Unfortunately, fibroblast cells were not available for P2.

### Patient fibroblasts show evidence of mitochondrial dysfunction

Given the integral role of mitochondria in ATP production, we first assessed oxygen consumption in patient fibroblasts, as this is a key indicator of changes to mitochondrial function. Compared to control cells, we observed marked decreases in both basal (around half) and maximal (around two-third) oxygen consumption (Figure 2a), as well as spare respiratory capacity (Figure 2b). These findings are indicative of severely reduced mitochondrial respiration in patient fibroblasts, consistent with an underlying mitochondrial dysfunction contributing to the patient phenotype.

**Figure 2:**
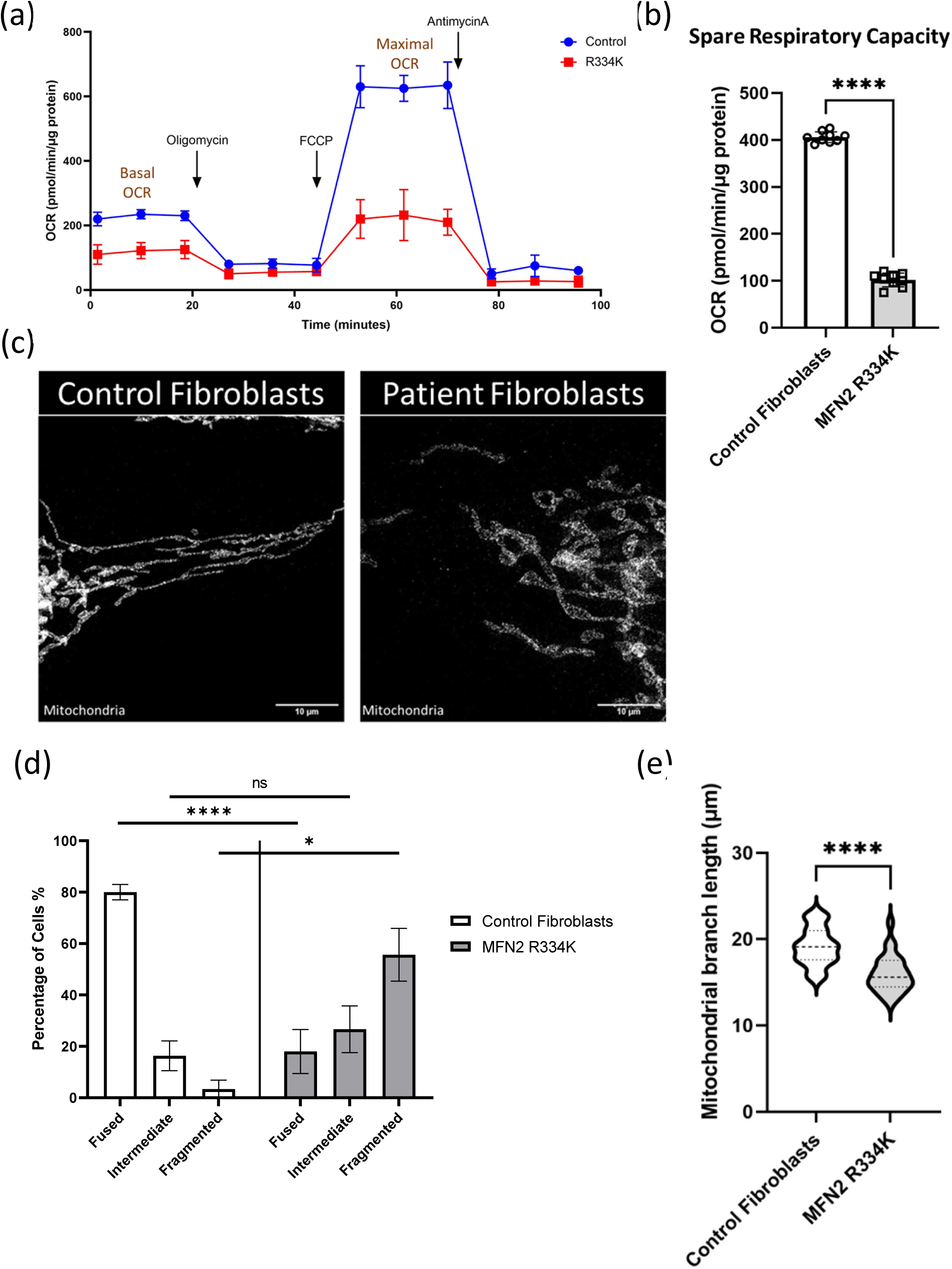
MFN2 R334K leads to severe mitochondrial dysfunction: (a) SeaHorse Oxygen Consumption Rate (OCR) indicating control and patient fibroblasts, measured using XF23 extracellular flux analyzer (2 biological replicates with 12 technical replicates). The error bars on the graph indicate the mean OCR +/ SD. (b) Quantification of spare respiratory capacity in control fibroblasts and patient fibroblasts (2 biological replicates with 12 technical replicates); the bars indicate mean +/- SD, statistics are from an unpaired t-test. (c) Representative STED confocal microscopy images showing mitochondrial network morphology in control vs MFN2 R334K patient fibroblasts, stained using anti-TOMM20. (d)Graphical representation of qualitative analysis of mitochondrial branch appearance (fused/intermediate/fragmented), from analysis of 150 images including 3 biological replicates, the bars indicate mean for each category +/ SD, statistics are from an unpaired t-test. (e) Violin plot showing quantitative analyses of mean mitochondrial branch length in control and patient fibroblasts (from 150 cells including 3 biological replicates), with violin plots indicating median and interquartile range (IQR), from an unpaired t-test. Statistical values indicate the following: ns P > 0.05, * P≤ 0.05, **** P ≤ 0.0001.

Next, we investigated the canonical function of MFN2 in mitochondrial fusion by visualizing the mitochondrial network after immunofluorescent labeling for the mitochondrial marker TOMM20 (Figure 2c). In comparison to the control fibroblasts, which showed elongated and reticular mitochondrial network morphology, patient fibroblasts qualitatively displayed more fragmented mitochondrial networks (Figure 2d). Quantitative analysis of the mitochondrial networks confirmed patient fibroblasts had shorter mitochondria (Figure 2e). Together, these findings are consistent with impaired mitochondrial fusion leading to more fragmented mitochondria.

### Changes in mitochondrial nucleoids and mtDNA copy number depletion in patient fibroblasts

To provide further evidence of impaired mitochondrial fusion, we investigated the distribution and abundance of mtDNA, which can be altered by impairments to mitochondrial fusion, are linked to loss of MFN2 function, and have been observed for some MFN2 variants [48, 56, 64]. Immunofluorescence imaging of mtDNA nucleoids (Figure 3a) revealed that the patient fibroblasts exhibited approximately half as many nucleoids as control fibroblasts (Figure 3b), and that patient mtDNA nucleoids were smaller (Figure 3c). Consistent with the reduced number of mtDNA nucleoids, we detected a 40% decrease in the mtDNA copy number in the patient fibroblasts via qPCR (Figure 3d). Finally, we also investigated the presence of mtDNA deletions, which are sometimes seen with MFN2 loss of function [64]. However, no mtDNA deletions were detected (Figure 3e).

**Figure 3:**
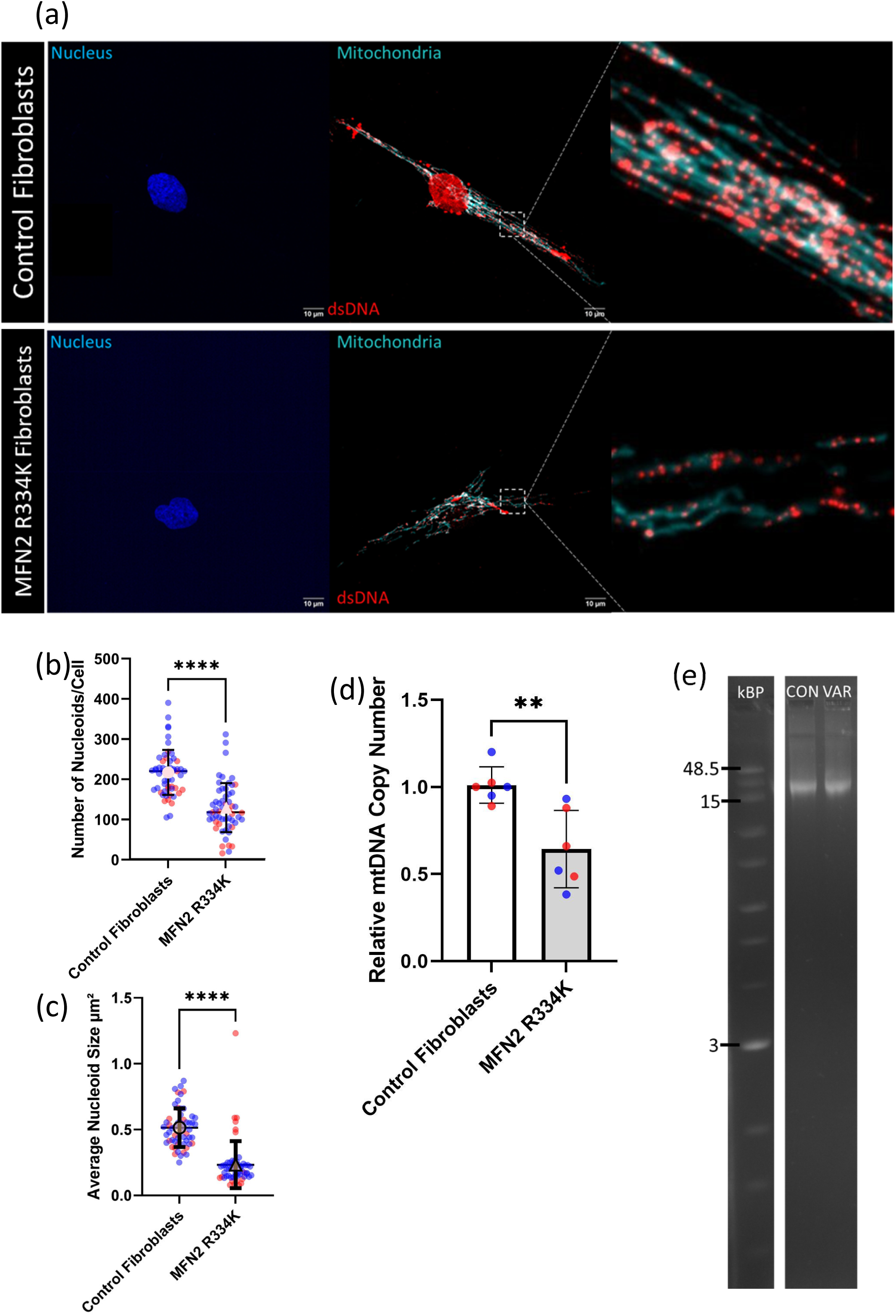
Alterations to mitochondrial DNA in patient fibroblasts. (a) Representative confocal images showing DAPI (nucleus) and mitochondrial nucleoids (anti-dsDNA) co-localized with the mitochondrial network (anti-TOMM20) for control (top panels) and patient fibroblasts (bottom panels). (b) Quantitative analyses of number of mitochondrial nucleoids per cell in control and patient fibroblasts, the colors indicate biological replicates, the lines indicate mean number of mito-nucleoids per cell +/ SD, statistics are from an unpaired t-test. (c) Quantitative analyses of average size(s) of mitochondrial nucleoids in control and patient fibroblasts, the colors indicate biological replicates, with the lines showing mean +/ SD, statistics are from an unpaired t-test. (d) Quantitative analysis of mtDNA copy number in control and patient fibroblasts, relative to 18S rDNA, the colors indicate biological replicates, bars indicate mean +/ SD, statistics are from an unpaired t-test. (e) Representative image of agarose gel to detect mtDNA deletions using 0.6% agarose gel following long-range PCR to target mtDNA amplification and ladder shown on the left. Statistics shown on the graph represent: ns P > 0.05, * P≤ 0.05, ** P ≤ 0.01, **** P ≤ 0.0001.

### Altered mito-ER contact sites and lipid droplet accumulation in patient fibroblasts

We investigated the size and abundance of mito-ER contact sites (MERCs) and lipid droplets, as these phenotypes can be altered in fibroblasts from patient harboring pathogenic MFN2 variants [30, 54]. As a proxy for MERCs, we utilized a proximity ligation assay (PLA) that generates a fluorescent signal when mitochondria and ER are within ∼40 nm (Figure 4a-b) [6]. Using confocal microscopy, we observed a decrease in the size and number of MERCs, which was confirmed by quantitative analyses (Figure 4c-d). We examined lipid droplets using the neutral lipid droplet dye BODIPY 493/503 (Figure 5a). Compared to control fibroblasts, we noted a distinct upregulation in the number (Figure 5b) and size (Figure 5c) of lipid droplet puncta in the R334K patient fibroblasts. These findings of decreased MERCs and increased lipid droplets are consistent with previously reported alterations in fibroblasts from patients with CMT2A harbouring MFN2 variants [9, 30].

**Figure 4:**
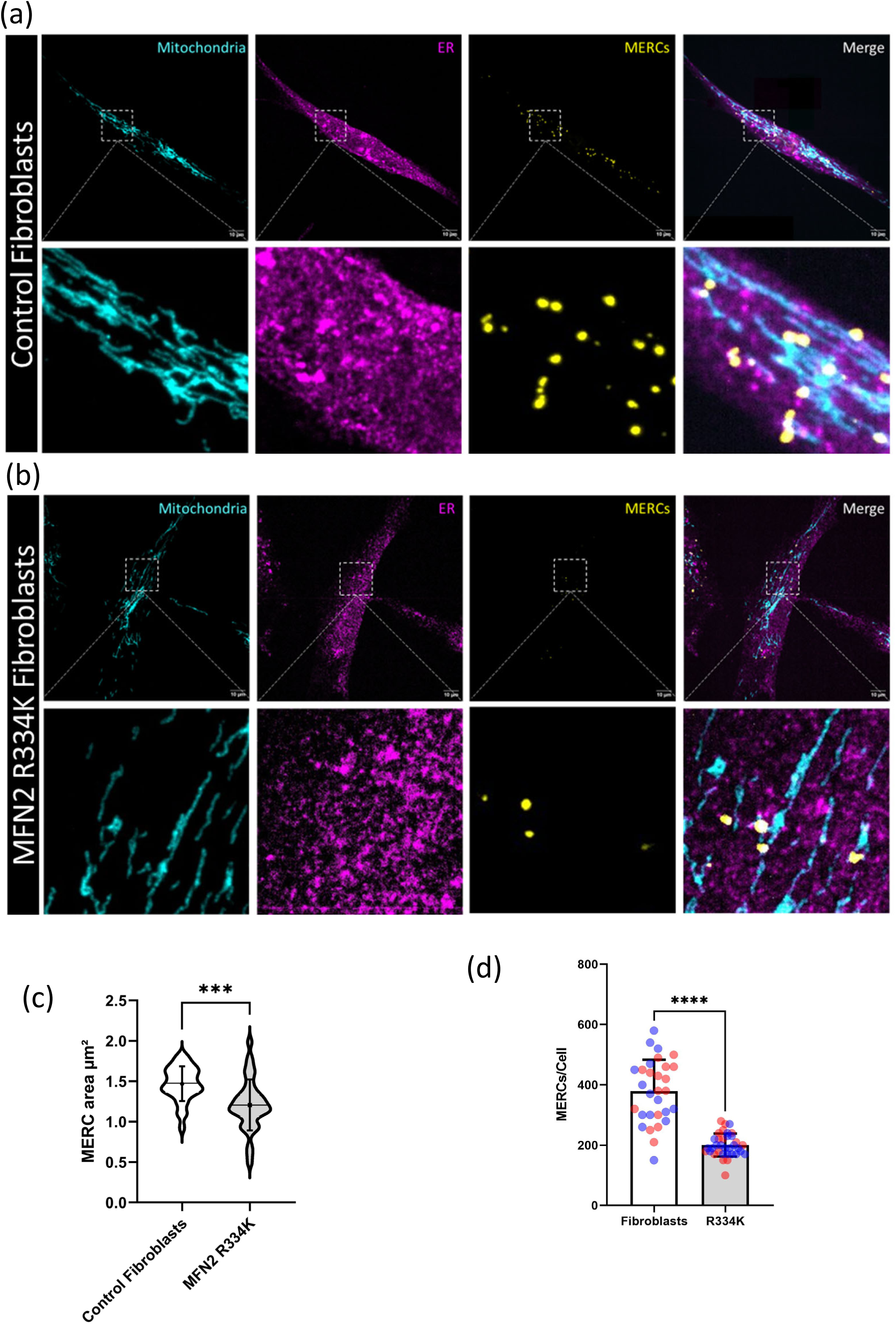
Changes to mito-ER contact sites in patient fibroblasts. (a) Top panel shows representative confocal images of mitochondria (anti-TOMM20) and ER (anti-calnexin) co-localization, MERCs (PLA probes) and merged images for control fibroblasts. The bottom panel shows zoomed representation of boxed region. (b) Top panel shows representative confocal images of mitochondria (anti-TOMM20) and ER (anti-calnexin) co-localization, MERCs (PLA probes) and merged images for patient fibroblasts. The bottom panel shows zoomed representation of boxed region. (c) Quantitative analyses of number of PLA signals (MERCs) per cell in control and patient fibroblasts, colors indicate biological replicates, the violin plots show distribution of data, while the lines indicate mean +/ SD, statistics are from an unpaired t-test. (d) Quantitative analyses of average size(s) of PLA puncta (size of MERCs) in control and patient fibroblasts bars indicate the mean size of PLA puncta +/ SD, statistics are from an unpaired t-test. Statistics used in the graphs represent the following*** P ≤ 0.001, **** P ≤ 0.0001.

**Figure 5:**
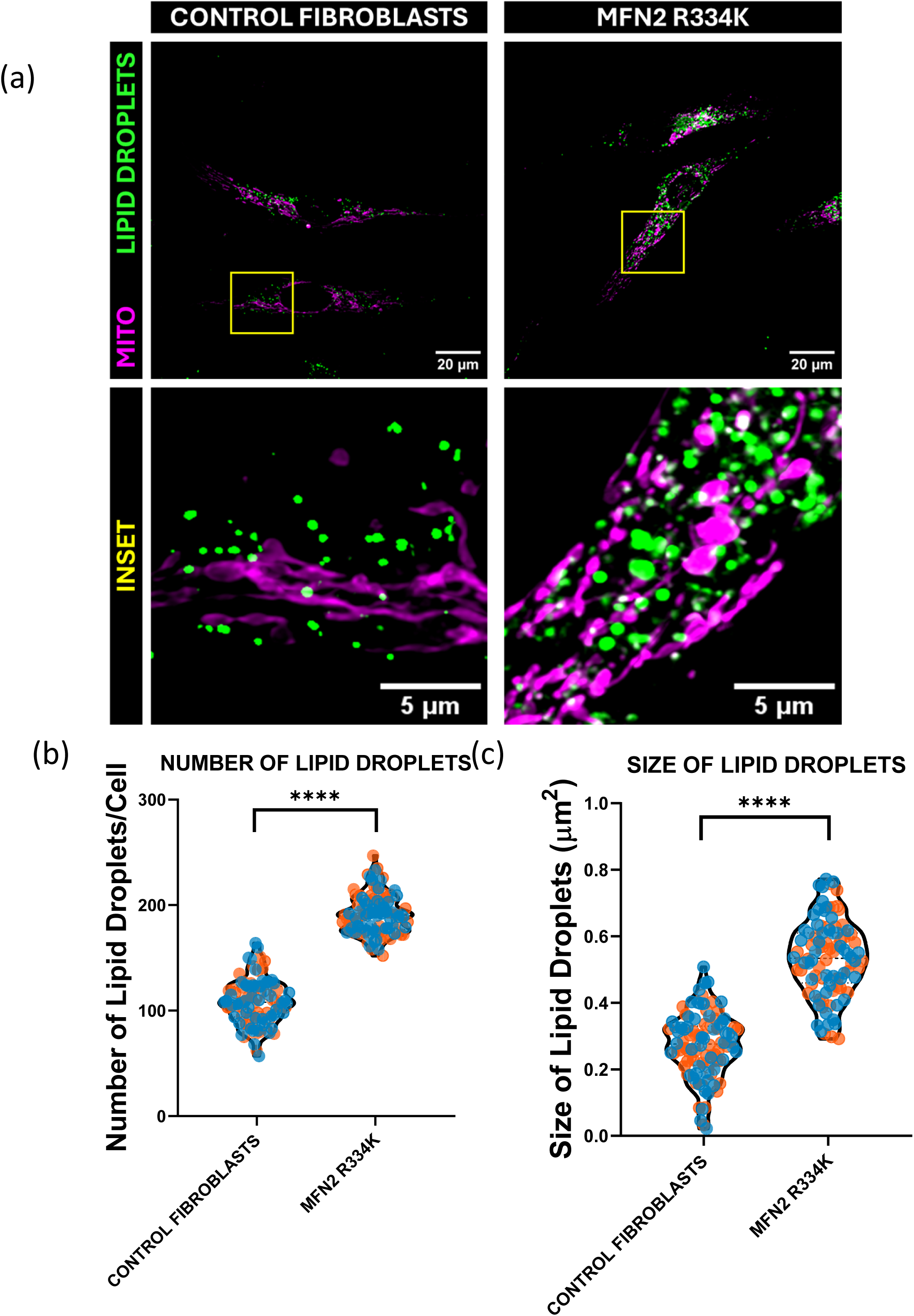
Changes to cellular lipid droplet accumulation in control and patient fibroblasts. (a) Representative confocal images from control (top) and patient (bottom) fibroblasts stained for lipid droplets using the neutral lipid droplet stain BODIPY™ 493/503. Quantification of average number (b) and (c) size of lipid droplets per cell in control and patient fibroblasts using BODIPY™ 493/503. The violin plots indicate median and interquartile ranges, colours indicate biological replicates and statistics are from an unpaired t-test. The statistics shown are represented by: **** P ≤ 0.0001.

### MFN2 and mitochondrial protein expression

As our findings were consistent with MFN2 dysfunction, we assessed if expression of MFN2 and other mitochondrial proteins was impacted. While we did not observe changes in MFN2 protein levels from total cell lysate (Figure 6a-b), we did observe a significant (∼50%) decrease in expression of the nuclear DNA encoded mitochondrial complex I subunit NDUFB8 (Figure 6a/c). Notably, there was no consistent significant change to the levels of the other OXPHOS proteins investigated here, including mtDNA-encoded MTCO1. The cause for this discrepancy in OxPhos protein expression is unknown, but likely does not reflect the mtDNA depletion, which would be expected to cause a global decrease in OxPhos protein expression as mtDNA encodes essential proteins for all the electron transport chain complexes except for complex II.

**Figure 6:**
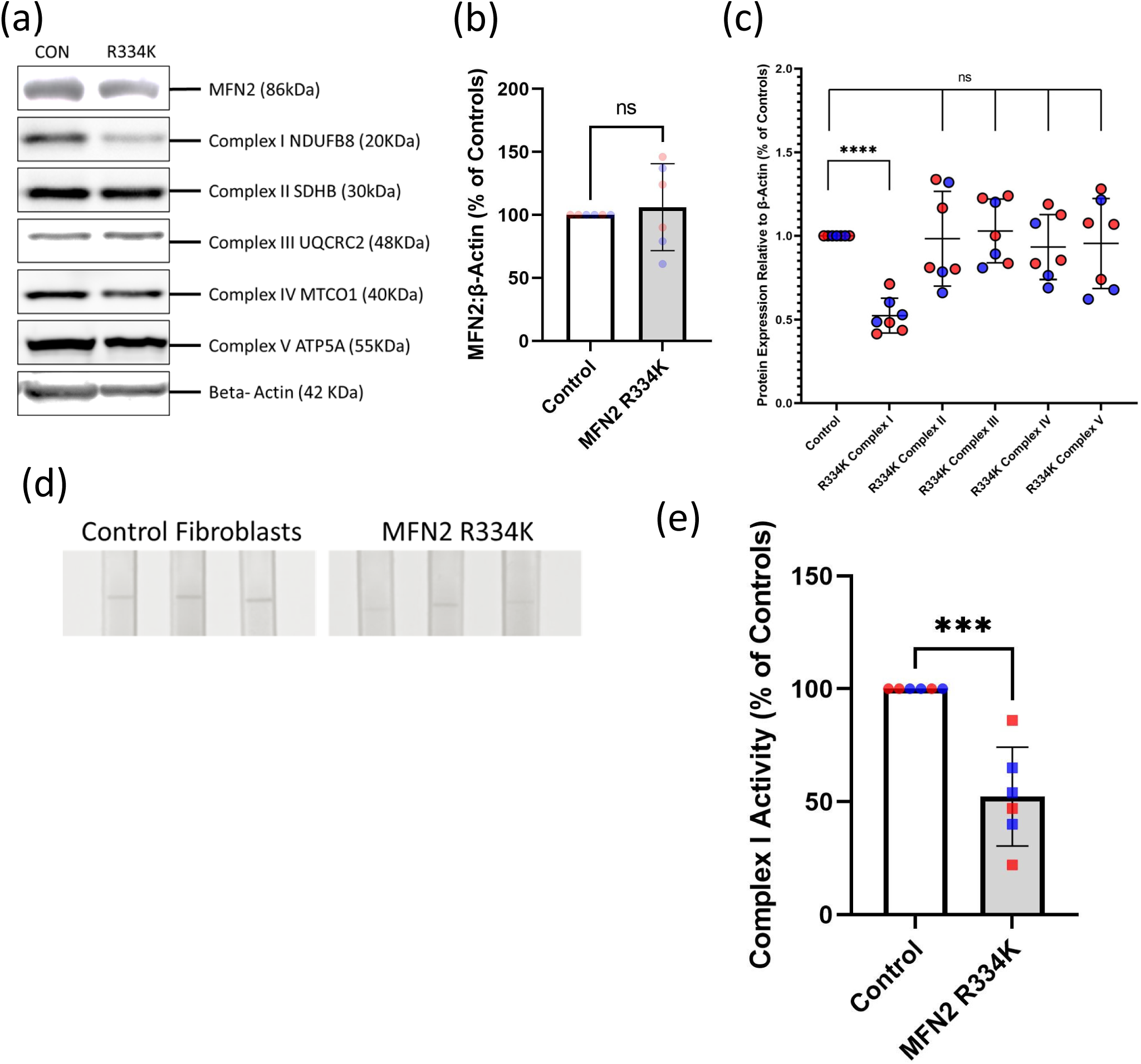
MFN2 R334K impedes levels and activity of OXPHOS Complex-I. (a) Representative Western Blot images from control and MFN2 R334K patient fibroblasts showing protein levels of MFN2 and the OxPhos complexes, as well as loading control Beta-Actin and mitochondrial loading control HSP-60. (b) Quantification of MFN2 levels in control fibroblasts and patient fibroblasts relative to Beta-Actin, colours indicate biological replicates, the bars indicate mean levels of MFN2 +/ SD, statistics show an unpaired t-test. (c) Quantification of mitochondrial OxPhos complexes (I-V) levels from Western Blot analyses in control and patient fibroblasts normalized to cellular beta-actin levels, colours indicate biological replicates, the lines indicate mean for each category mean +/ SD, statistics are from individual unpaired t-tests versus the control. (d) Representative images showing dipsticks used to characterize relative complex I activity in control and patient fibroblasts, using the complex I dipstick assay. (e) Quantification of relative complex-I activity performed using complex-I dipstick assay in control and patient fibroblasts, colours indicate biological replicates, while bars indicate mean for each category +/ SD, Statistics shown indicate: ns P > 0.05, *** P ≤ 0.001, **** P ≤ 0.0001.

### Complex I activity

Given the specific decrease in a Complex I protein, which is not typical of MFN2 dysfunction and likely not explained by mtDNA depletion, we wondered if the identified FOXRED1 missense variant might also have functional consequences that could contribute to the patient phenotype. As pathogenic defects in FOXRED1 are linked to dysfunction in complex I assembly and stability, we also examined Complex I activity. Consistent with the reduced protein expression, and possible complex I instability, we observed an ∼50% reduction in Complex I activity in patient fibroblasts (Figure 6d-e).

### In vitro re-expression of MFN2 R334K recapitulates Complex-I deficiencies

To test if the reduced Complex I activity in patient fibroblasts can be explained by the MFN2 R334K variant alone, we used a knockout re-expression system to test the pathogenicity of the R334K MFN2 variant in a different genetic background. Using U2OS MFN2 KO cells, we re-expressed either the WT MFN2 protein or the MFN2 R334K variant by viral transduction, selecting cells expressing endogenous levels of MFN2 expression (Figure 7c-d). To confirm the system recapitulates features of patient fibroblasts, we first looked at mitochondrial morphology. As expected, MFN2 KO cells had severely fragmented mitochondrial networks, which were rescued by re-expression of the WT protein in KO cells (Figure 7a-b). In line with the mitochondrial fragmentation observed in patient fibroblasts, we found that MFN2 KO cells re-expressing MFN2 R334K show partial rescue compared to MFN2 KO cells, but remained more fragmented than control cells or KO cells re-expressing WT MFN2. Next, we examined the expression of OxPhos proteins and Complex I function. MFN2 KO cells had severe reductions in Complexes I, II, III, and IV subunits, which were fully rescued by re-expressing WT MFN2. However, although Complex II, III and IV protein expression was rescued in MFN2 KO cells re-expressing the R334K variant, Complex I protein expression was reduced by ∼50% compared to controls (Figure 7c-d). Looking at Complex I activity, MFN2 KO cells showed a marked reduction that was fully rescued by WT MFN2. Although cells re-expressing MFN2 R334K showed a partial rescue compared to the KO, there was still a significant 50% decrease compared to controls (Figure 7e-f). Taken together, these findings confirm previous work showing MFN2 loss of function can lead to reduced Complex I activity [34, 45], and demonstrate that the MFN2 R334K variant alone can explain the 50% decrease in Complex I activity observed in patient fibroblasts.

**Figure 7.**
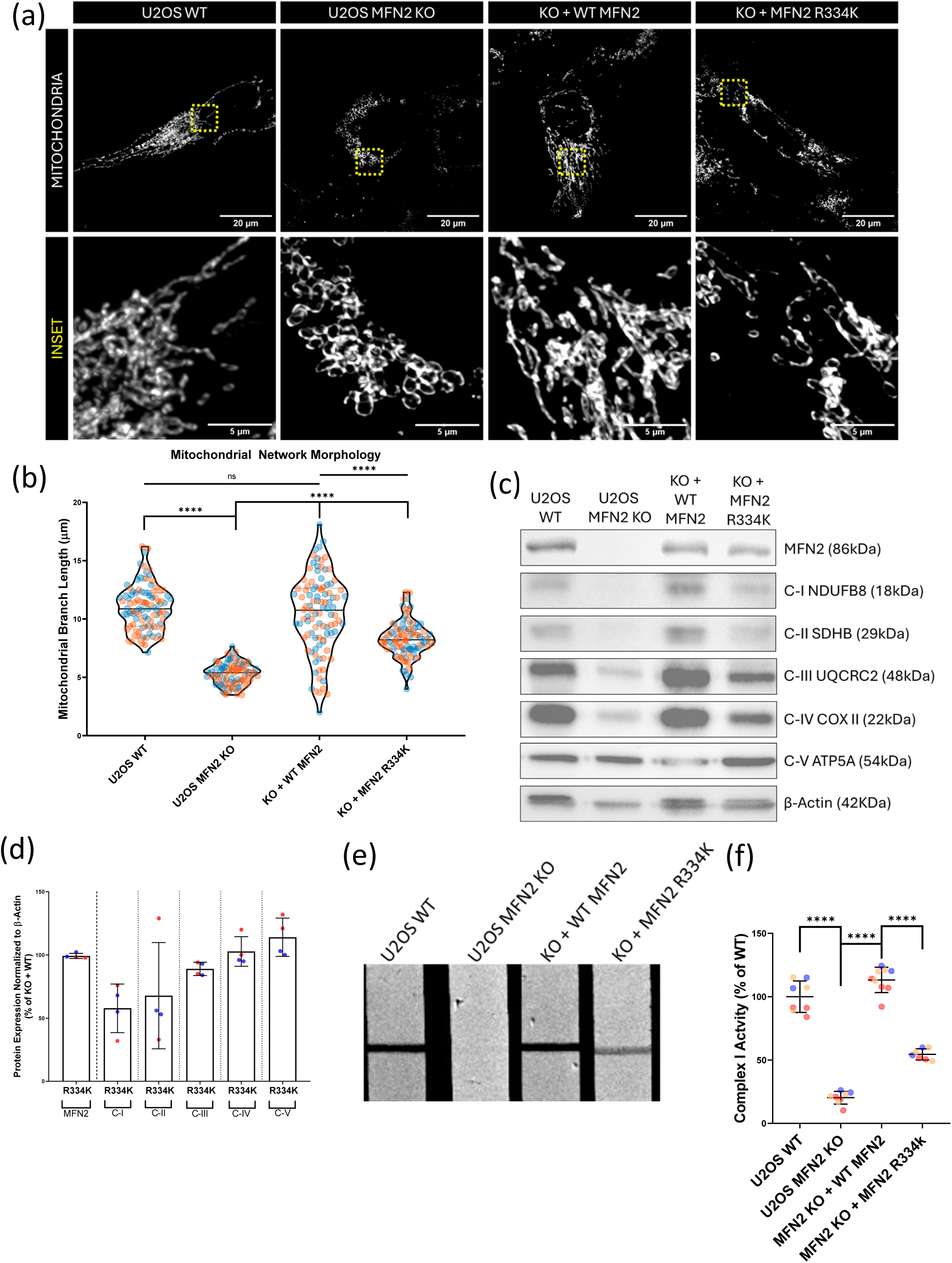
In vitro modelling of MFN2 R334K shows failure to rescue mitochondrial fragmentation and complex-I deficiencies. (a) Representative confocal images showing mitochondrial network morphology, labelled using anti-TOMM20 antibody for WT U2OS, U2OS MFN2 KO and re-expression of WT MFN2 or MFN2 R334K in KO cells. (b) Quantitative analyses of mitochondrial network morphology represented by mean mitochondrial branch length, colours indicate biological replicates, violins plots show the median branch length and inter-quartile range, statistics were performed using a one-way ANOVA. (c) Representative western blot images showing re-expression levels of MFN2 (WT/R334K) and corresponding levels of OXPHOS complex subunits, as well as loading control beta-actin. (d) Quantitative analyses in WT U2OS/MFN2 KO/WT re-expression/MFN2 R334K re-expression in KO cells showing levels of proteins (MFN2 re-expression levels and OXPHOS complexes); the colours indicate biological replicates, bars show the mean protein levels +/- SD, statistics are from an unpaired t-test. (e-f) Complex-I dysfunction in U2OS MFN2 R334K re-expression cells with (e) representative images showing complex-I activity dipsticks and (f) Quantitative analyses of complex-I dipstick assay, the colours indicate biological replicates, the lines show mean complex I activity +/- SD, statistics are from one-way ANOVA. The statistics represent the following: ns P > 0.05, ** P ≤ 0.01, **** P ≤ 0.0001.

### In vivo modelling of MFN2 R334K in Drosophila melanogaster

To model the pathogenicity of the R334K MFN2 variant in vivo, we used a knockdown re-expression paradigm in Drosophila (Figure 8a) that has been used to study the function of other human MFN2 variants [19]. In this approach, depleting the MFN2 orthologue in flies, marf, can be rescued by re-expressing WT human MFN2 (hMFN2). First, we examined the effect of whole-body knockdown/re-expression using three different driver lines: Actin (act5c-Gal4), Tubulin (tub-Gal4) or Daughterless (da-Gal4). Whole-body knockdown of marf was larval lethal for all three promoters tested. While act5c-Gal4 and tub-Gal4 showed no survival, the weaker da-Gal4 driver had ∼10% survival (Figure 8b). Meanwhile, re-expression of WT hMFN2 rescued survival to ∼60% (Figure 8b), confirming the ability of the WT human MFN2 to function in flies. In contrast, re-expression of MFN2 R334K was insufficient to rescue the larval lethality (Figure 8b) in any of the whole-body driver lines.

**Figure 8:**
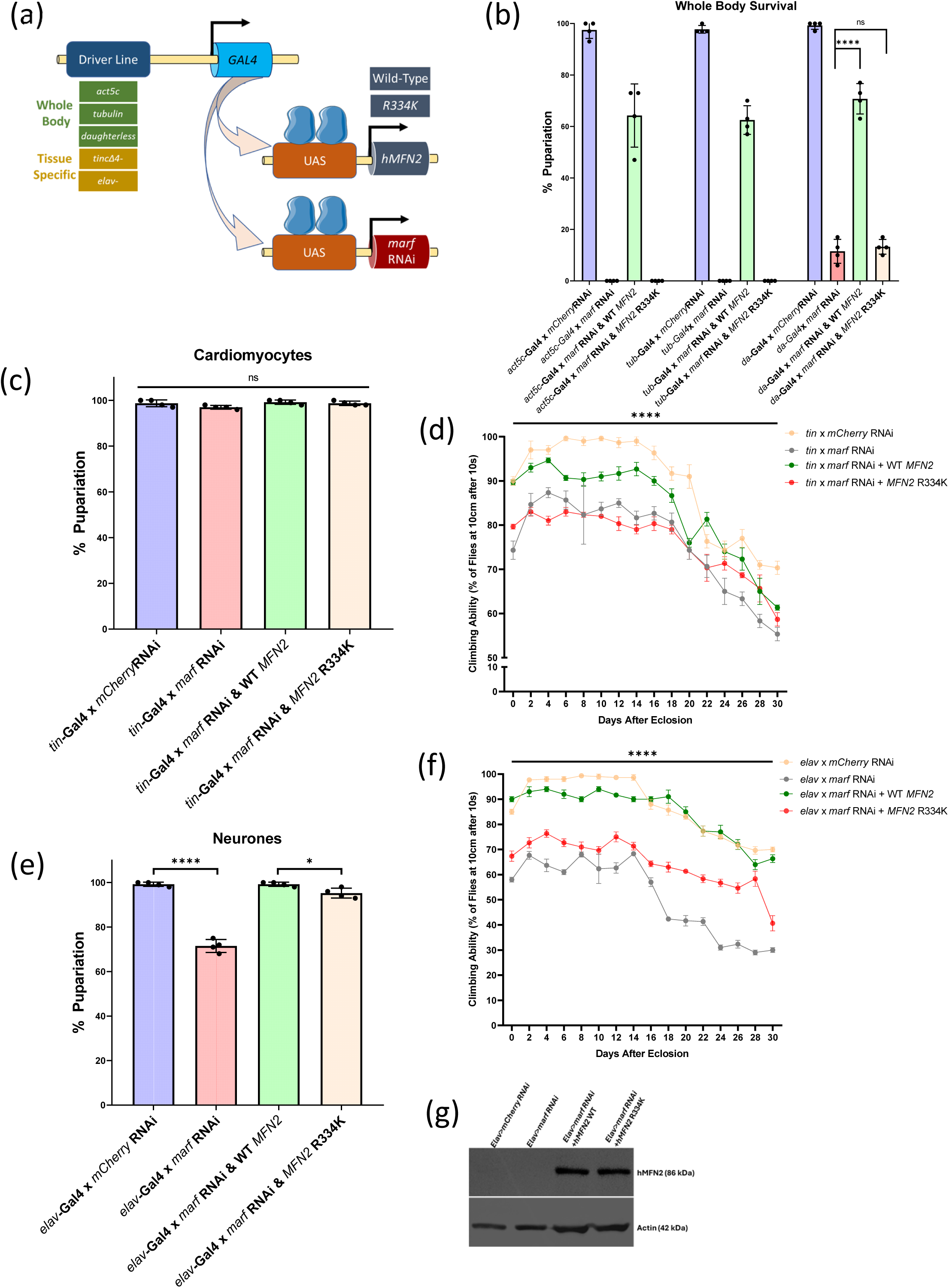
In vivo analyses of MFN2 R334K using Drosophila melanogaster show lethality and locomotor dysfunctions. (a) Schematic representing Gal4-UAS approach for human disease modelling in Drosophila melanogaster, with Gal4 under the control of whole body/tissue specific promoters leading to expression of UAS-hMFN2 on chromosome II (phiC31 integrase approach) and UAS-marf RNAi on chromosome III. UAS-mCherry RNAi was used as a control for the Gal4-UAS system. (b) Whole-body survival assay in control, marf knockdown and re-expression of hMFN2 WT and MFN2 R334K fly lines using Actin (act5c-Gal4), Tubulin (tub-Gal4) and Daughterless (da-Gal4) driver lines. n=80 flies, with four independent replicates. Bars show mean survivability +/- SD, statistics are from unpaired t-tests. (c) Tissue-specific survival assay for cardiomyocytes, using control, marf knockdown and re-expression of hMFN2 WT and MFN2 R334K fly lines using tinman (tin-Gal4) driver line. n=80, with four independent replicates; The bars on the graph show mean survival +/- SD, statistics are from unpaired t-tests. (d) Negative geotaxis assay to look for climbing ability of flies with changes to cardiomyocytes (tin) knockdown and re-expression of human MFN2, lines show mean +/- SD, statistics are from unpaired t-tests at each time point for WT-re=expression vs R334K re-expression. (e) Tissue-specific survival assay for neurons, using control, marf knockdown and re-expression of hMFN2 WT and MFN2 R334K fly lines using elav (elav-Gal4) driver line. n=80, with four independent replicates; The bars on the graph show mean survival +/- SD, statistics are from unpaired t-tests. (f) Negative geotaxis assay to look for climbing ability of flies with changes to neurons (elav) knockdown and re-expression of human MFN2, lines show mean +/- SD, statistics are from unpaired t-tests at each time point for WT-re-expression vs R334K re-expression. (f)Representative Western Blot images showing expression of human MFN2 protein expression in in control, marf knockdown and re-expression of hMFN2 WT and MFN2 R334K fly lines under the control of elav-Gal4. (b) Statistics represent the followingns P > 0.05, * P≤ 0.05, ** P ≤ 0.01, **** P ≤ 0.0001.

Next, given the larval lethality of the whole-body marf knockdown and MFN2 R334K re-expression, we used a tissue-specific drivers to examine the effects in neurons (elav-Gal4) and cardiomyocytes (tin-Gal4), which are relevant to the neuromuscular patient pathology. Knockdown of marf in cardiomyocytes showed no significant lethality (Figure 8c). Next, we assayed for locomotor function using a negative geotaxis climbing assay. We observed a marked decrease in climbing ability of flies with knockdown of marf, which is nearly completely rescued by re-expressing WT MFN2, but not by re-expression of MFN2 R334K (Figure 8d). Using the neuronal driver, we saw 70% survival with marf knockdown, which was completely rescued with WT MFN2, while MFN2 R334K had 95% survival (Figure 8e). In terms of climbing ability with the neuron driver line marf knockdown also led to a significant decrease, which was completely rescued by WT MFN2 climbing. In contrast, MFN2 R334K provided only a slight rescue (Figure 8f). Importantly, we confirmed similar levels of WT MFN2 and MFN2 R334K in our fly lines (Figure 8g). Overall, these data show that the MFN2 R334K variant has reduced functionality and is unable to rescue marf knockdown in vivo to the same level as WT MFN2. Moreover, the deficits seen with neuronal and cardiomyocyte-specific knockdown, reflect the severe pathology seen in the patient.

### Pharmacologic ISR activation rescues MFN2 R334K dysfunction in patient cells

Given our previous work showing ISR activation rescues mitochondrial fragmentation and oxygen consumption in MFN2-deficient cells [5, 8], we speculated that ISR activation would ameliorate dysfunction in MFN2 R334K patient fibroblasts. To this end, a 6-hour treatment with three distinct ISR activators (Halofuginone, 3610, and Parogrelil) completely rescued mitochondrial morphology in R334K patient fibroblasts (Figure 9a-b/Supplemental Figure 1a-d). Furthermore, all three ISR activators also partially rescued OCR deficits (Figure 9c). Importantly, co-treatment with the ISR inhibitor ISRIB prevented the rescue of both morphology and OCR for all three compounds, confirming that this rescue is dependent on ISR activation, rather than any potential off-target effects of the drugs. Overall, these findings show that pharmacological ISR activation is beneficial for MFN2 dysfunction, even for severe perturbations such as those caused by MFN2 R334K.

**Figure 9:**
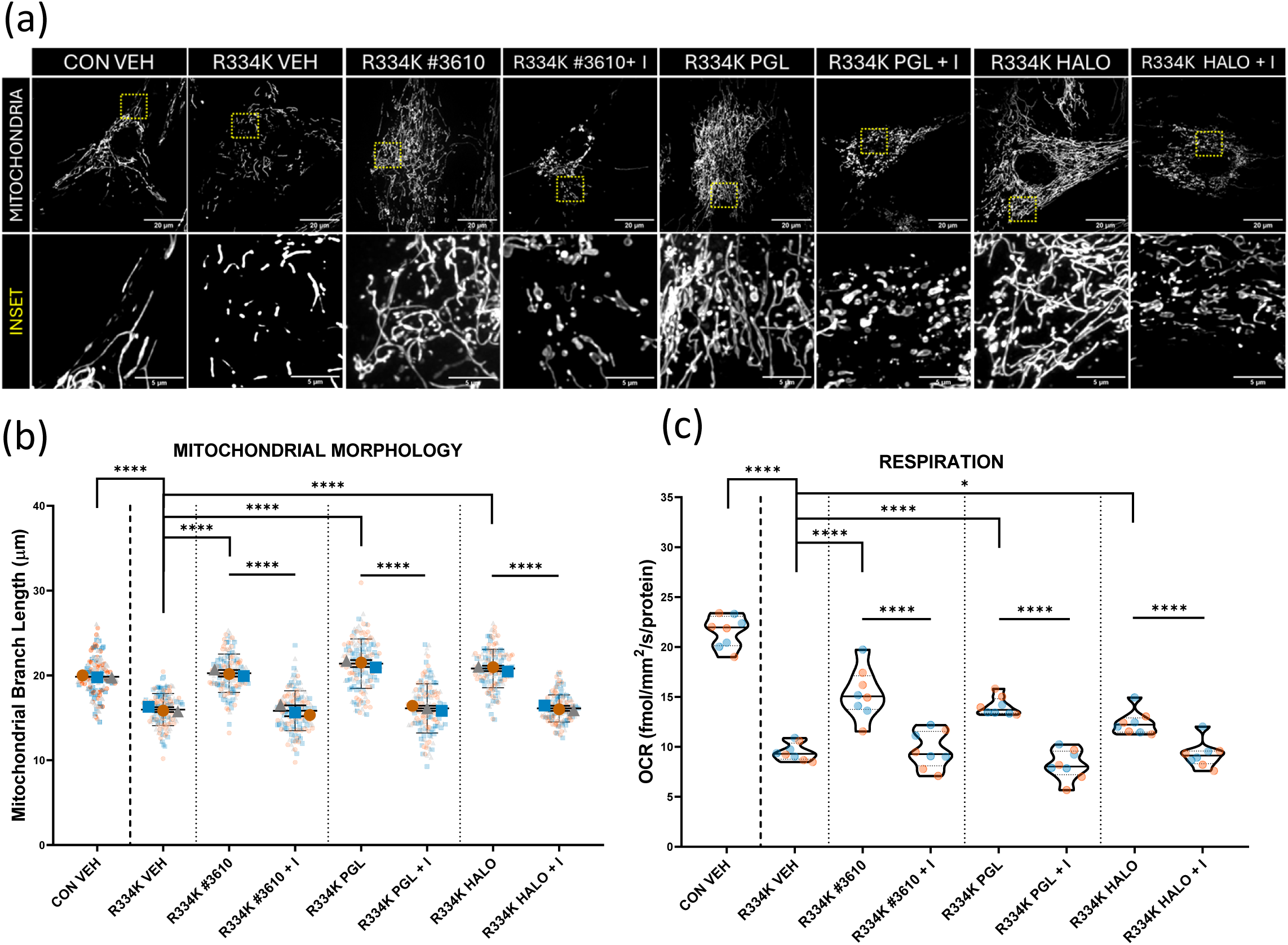
ISR activation rescues mitochondrial fragmentation and boosts cellular oxygen consumption in MFN2 R334K patient cells. (a) Representative confocal images showing mitochondrial network morphology using MitoTracker green for control fibroblasts vs MFN2 R334K patient fibroblasts treated with ISR activators/inhibitors for 6 hours with vehicle, #3610 (10uM), Parogrelil (10uM) and Halofuginone (100nM), along with ISRIB (200nM). (b) Quantitative analyses of mitochondrial network morphology for control/patient fibroblasts treated with ISR activators, points on the graph indicate biological replicates, lines show mean mitochondrial branch length +/- SD, statistics come from one-way ANOVA. (c) Quantitative analyses of cellular oxygen consumption rate measured using Resipher OCR, for control fibroblasts and patient fibroblasts treated for 6 hours with vehicle, #3610 (10uM), Parogrelil (10uM) and Halofuginone (100nM), along with ISRIB (200nM). Colours indicate biological replicates, violin plots show the median OCR and IQR, one-way ANOVA. Statistics on the graphs represent the following: ns P > 0.05, * P≤ 0.05, **** P ≤ 0.0001.

## Discussion

We report three siblings who presented with severe neonatal encephalopathy, hypotonia, areflexia and arthrogryposis and all three were homozygous for a novel missense variant in MFN2 p.(Arg334Lys). We speculate that the location of the R334K MFN2 variant is crucial for proper protein function, possibly by altering the nearby GTPase domain of the protein. Though CMT2A can be an early-onset disease that can eventually restrict patient movement as the disease progresses, the patients reported here exhibited lack of movement at birth, indicative of a much more severe neonatal onset. Moreover, all three patients succumbed in infancy (<6 weeks) and we would consider this presentation as being neonatal lethal with ventilation. No improvement was observed over their clinical courses. Although areflexia and hypotonia phenotypes are not typical of CMT2A, there are a few previous reports of both phenotypes in patients harbouring MFN2 variants with severe early-onset presentation [35, 43, 48, 51, 57, 65]. However, the patients we report here presented with symptoms immediately upon birth, as opposed to the examples in the previous reports, where symptoms presented from 6-12 months. With respect to the early onset of severe phenotypes linked to MFN2 dysfunction, there is a recent report of a patient with severe CNS complications who harboured a homozygous 50 amino acid deletion in MFN2 with antenatal presentation that resulted in pregnancy termination at 31 weeks [13]. In this regard, the fact that loss of MFN2 function leads to embryonic lethality in mice [12] and larval lethality in fruit flies [19], is consistent with a severe loss of MFN2 function due to the R334K variant.

In support of the notion that the R334K variant impairs MFN2 functions, we observed changes across all the functions we explored, including mitochondrial network morphology, oxidative phosphorylation, complex I activity, MERCs, mtDNA copy number and nucleoids, as well as cellular lipid droplets. These findings suggest that the novel R334K variant causes severe dysfunction of the MFN2 protein. To our knowledge, there are no previously characterized MFN2 CMT2a variants with defects across all these functions [64]. However, to some degree this finding likely reflects the fact that few MFN2 variants have been characterized extensively for their impact on MFN2 functions, highlighting our limited understanding of how MNF2 variants impact MFN2 functions. In terms of MFN2 variants associated with more severe cellular phenotypes, it is notable that MFN2 variants that correlate with reduced mtDNA copy number and impaired oxidative phosphorylation are reported in patients that tend to present with earlier-onset and severe clinical outcomes [33, 54, 60]. Collectively, this evidence is consistent with severe MFN2 dysfunction contributing to the very early onset of clinical symptoms in the patients we report here.

Considering the additional homozygous FOXRED1 variant in P1, it is also worth briefly discussing the 50% Complex I deficiency in this patient’s fibroblasts, which is not typical of pathogenic MFN2 variants. Notably, complete loss of MFN2 function can lead to a specific deficit in Complex I expression and function [34], which we also observe in U2OS KO cells. Moreover, Complex I dysfunction was also reported with the MFN2 variant harbouring a 50 amino acid deletion resulting in pregnancy termination [13]. Thus, MFN2 dysfunction alone is sufficient to cause Complex I deficiency. Although FOXRED1 is a molecular chaperone important for the assembly of mitochondrial complex I [22, 42], several lines of evidence argue against a role for the FOXRED1 I485N the patients’ phenotype. First, the patient phenotypes do not match. Homozygous or compound heterozygous recessive FOXRED1 variants cause mitochondrial complex I deficiency, nuclear type 19 (MC1DN19), where only a handful of cases have been described [4, 11, 20, 22, 24, 27, 66]. Clinical phenotypes of MC1DN19 mimic common mitochondrial disorders such as Leigh syndrome and mitochondrial encephalopathy [11, 27, 36], including lactic acidosis and growth restrictions. In contrast, there was no evidence of lactic acidosis in these patients. Meanwhile, though MC1DN19 patients can have hypotonia [20, 27], and can also be diagnosed with restricted movement accompanying Leigh syndrome [22], there are no reports of areflexia in FOXRED1 patients, who typically survive well past infancy [4]. Secondly, the genetics also argue against I485N FOXRED1 variant contributing to pathology, as FOXRED1 is autosomal recessive, while P2 was heterozygous for the I485N FOXRED1 variant. Thus, despite the fact that novel I485N FOXRED1 variant reported here is located at the C-terminus of the protein, similar to other pathogenic FOXRED1 variants described to date, and the fact that FOXRED1 patient fibroblasts also have reduced Complex I activity [20] (with patient myoblasts reported with deficiencies in both Complex I and Complex II [66]), this variant was ruled out as a contributing factor for the phenotype of P1 and P2, who had identical presentations. Finally, our results in MFN2 KO cells re-expressing the R334K variant show that this variant alone is sufficient to cause a 50% decrease in Complex I activity, fully explaining the loss of Complex I activity in patient fibroblasts.

As there is no cure for CMT2A or other MFN2-mediated pathology, it is notable that pharmacological ISR activation ameliorates the dysfunctions tied to mitochondrial fragmentation and impaired oxygen consumption in MFN2 R334K patient cells. Notably, one of the ISR activators we tested, Parogrelil, is an FDA-approved drug that has passed phase I clinical trials [26, 28]. However, before clinical use for MFN2 pathology, further work is needed to decipher mechanisms of ISR-dependent rescue, benefits beyond cultured cells, as well as temporal and dosage effects. Nonetheless, ISR activation is a promising therapeutic intervention for MFN2-mediated pathology.

Rapid whole exome sequencing (WES) has repeatedly shown that a quicker time to diagnosis guides goals of clinical care, including less failed treatments, and ultimately lowers the financial burden on the health care system [14, 23, 37]. While previous reports suggests that between 30-50% of WES cases are solved or partly solved by a known disease-associated gene, additional cases may reveal novel genes of uncertain significance (GUS). This study demonstrates the utility of rapid WES accompanied by functional experiments to assess these GUS and provide a diagnosis. Functional studies also have the potential to discover new treatment options for novel genes. However, as this study demonstrates, this is dependent upon the damaging effects of the variant on the normal function of the gene and/or protein. Regardless, this study demonstrates the value of collaboration between clinical and research colleagues.

While we acknowledge the limitations of studying a novel protein variant in a single family, our functional studies in cells (both in patient fibroblasts and MFN2 KO cells re-expressing the R334K variant), and in an animal model clearly demonstrate that functional impairment of the R334K MFN2 variant is sufficient to explain the observed cellular and patient phenotypes. The fact that that U2OS cells re-expressing the R334K variant recapitulates the key cellular phenotypes of mitochondrial fragmentation and reduced oxygen consumption show that this variant, rather than the FOXRED1 variant or some other undetected variant, is responsible for the observed mitochondrial dysfunction in patient fibroblasts. Meanwhile, modeling of the R334K variant in Drosophila provides in vivo evidence that the R334K variant can explain the severe early onset patient phenotypes. Thus, combined with clear genetic evidence, this functional work strongly argues for R334K as a novel pathogenic MFN2 variant causing a severe early onset pathology. This work also highlights the fact that MFN2 dysfunction can cause much more severe pathology than CMT2A typically recognized to be associated with MFN2 variants.

## Data Availability

All data produced in the present work are contained in the manuscript

## Acknowledgements

The authors would like to thank the study participants and their family. We would also like to thank Matthew Lines for his input on the manuscript. We would like to thank Dr. Paul Marcogliese (University of Manitoba, Canada) for his expertise and technical guidance with the Drosophila work.

## Author contributions

Designed and/or performed and analyzed mitochondrial or Drosophila experiments, MZ, CC, SG, RLW and TES; Clinical evaluation of patients and write up, JLM, BM, AMI, FPB; Analyzed exome data AVP; Drafted the manuscript or figures, MZ, AVP, BM, TES. All authors discussed the results and commented on the manuscript.

## Figure Legends

Supplemental Figure S1: ISR activation rescues mitochondrial fragmentation and boosts cellular oxygen consumption. (A) Representative confocal images showing mitochondrial network morphology using MitoTracker green for control fibroblasts treated with the following ISR activators/inhibitors for 6 hours with vehicle, #3610 (10uM), Parogrelil (10uM) and Halofuginone (100nM), along with ISRIB (200nM). (b) Quantitative analyses of mitochondrial network morphology for control fibroblasts treated with ISR activators, points indicate biological replicates, bars show mean +/- SD, statistics represent a one-way ANOVA. (c) Representative confocal images showing mitochondrial network morphology using MitoTracker green for MFN2 R334K patient fibroblasts treated with ISR activators/inhibitors for 6 hours with vehicle, #3610 (10uM), Parogrelil (10uM) and Halofuginone (100nM), along with ISRIB (200nM). (d) Quantitative analyses of mitochondrial network morphology for patient fibroblasts, colours indicate biological replicates, symbols indicate means of biological replicates, lines show mean +/- SD, one-way ANOVA. (e) Quantitative analyses of cellular oxygen consumption rate measured using Resipher OCR, for control fibroblasts and patient fibroblasts treated for 6 hours with vehicle, #3610 (10uM), Parogrelil (10uM) and Halofuginone (100nM), along with ISRIB (200nM). Colours indicate biological replicates, violin plots show median and IQR, statistics represent a one-way ANOVA. Statistical symbols represent ns P > 0.05, * P≤ 0.05, **** P ≤ 0.0001.

