## Supplementary figures and images for "A novel variant in MFN2 linked to a lethal disorder of neonatal onset"

### Supplemental Information

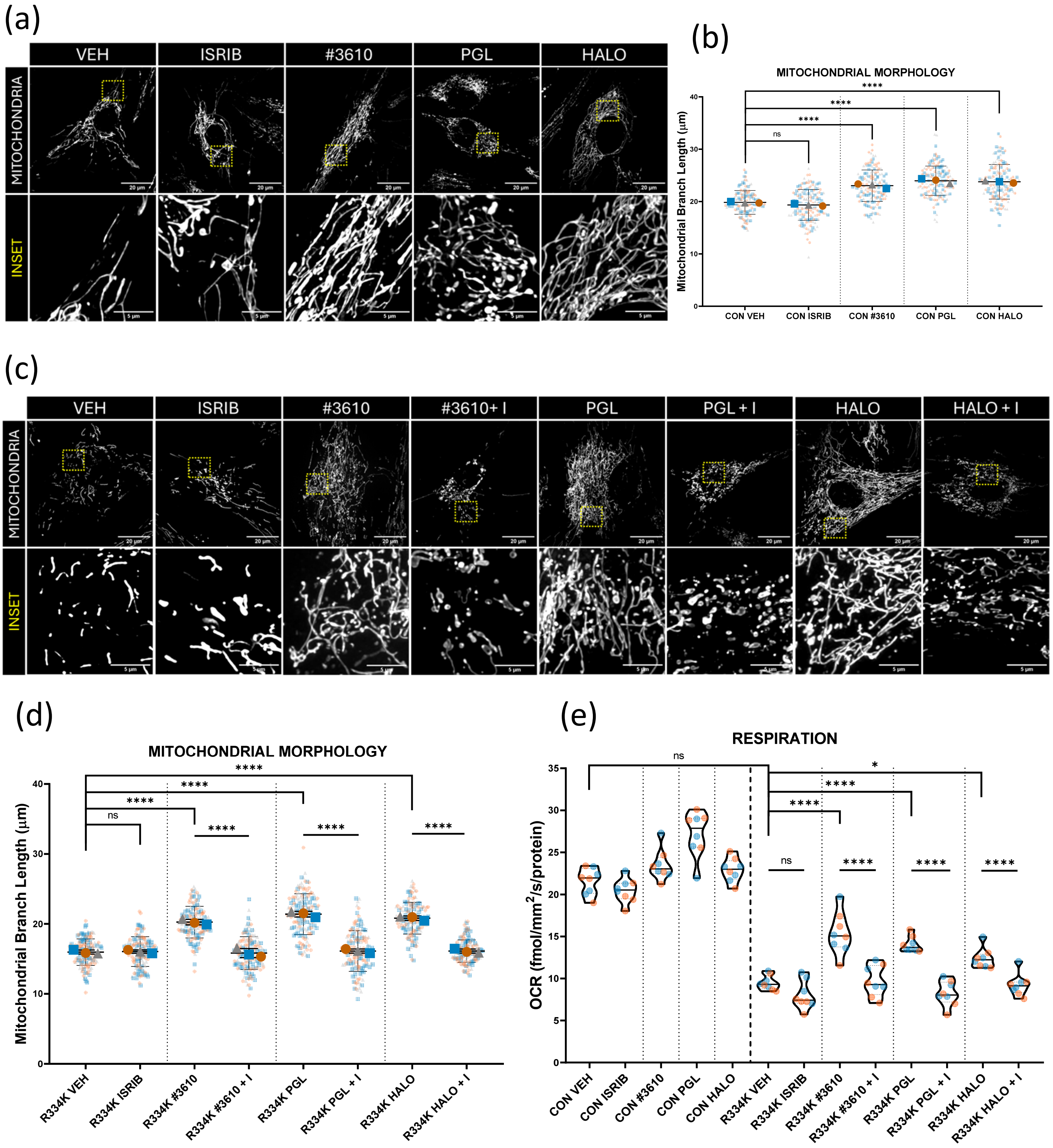


Supplemental Figure S1
